# Covid-19 Cases in India: A Visual Exploratory Data Analysis Model

**DOI:** 10.1101/2020.09.11.20193029

**Authors:** S Jayesh, Shilpa Sreedharan

**Affiliations:** Department of Mechanical Engineering, School of Engineering, Cochin university of Science and Technology, Kochi, Kerala-682022, email-; Medical Officer, Wellness Solutions, Kochi, Kerala

**Keywords:** Covid-19, India, data analysis, exploratory analysis

## Abstract

Covid-19 outbreak was first reported in Wuhan, China. The deadly virus spread not just the disease, but fear around the globe. On January 2020, WHO declared COVID-19 as a Public Health Emergency of International Concern (PHEIC). First case of Covid-19 in India was reported on January 30, 2020. By the time, India was prepared in fighting against the virus. India has taken various measures to tackle the situation. In this paper, an exploratory data analysis of Covid-19 cases in India is carried out. Data namely number of cases, testing done, Case Fatality ratio, Number of deaths, change in visits stringency index and measures taken by the government is used for modelling and visual exploratory data analysis.

## 1. Introduction

The new panic in town, the novel Coronavirus disease, abstracted as COVID-19 was first reported as an unexpected outbreak in Wuhan, the capital city of Hubei Province in the People’s Republic of China. Within a few weeks, the disease emerged contagious and spread to different parts of the world. The stupendous containment measures taken by the Chinese government to arrest the spread of the virus got washed down the drain. Being victims of the panic and lack of sustainability issues, countries around the globe went to lock down. A rising infectious disease alarms as rapid spreading, putting in danger the health of a large community, the weak being most vulnerable. Immediate action at the root level is required to tackle the virus [1–3]. The initial case of Covid-19 in India was reported on January 30, 2020 in Kerala, a southern state in India. The Kerala government accepting advices from the central boosted up its health and hygiene department and bought in vivid containment, isolation, tracking and many other containment measures to arrest the community spread. On 22^nd^ March 2020, India observed its first ever 14-hour controlled public curfew taking into consideration about the serious spread of the disease to restrict public movement. This was followed by nationwide lockdowns. On 24th March, the Prime Minister announced a nationwide lockdown for 21 days, distressing the entire 1.3 billion-person population of India. On 14th April, the nationwide lockdown continued extension and be in force till 3rd May, followed by 2 weeks extensions starting 3rd and 17th May with significant relaxations. However, starting with June, the government started unlocking procedure in three phases [4–6].

The United Nations (UN) and the World Health Organization (WHO)has particularly acknowledged India’s reaction to the pandemic as ‘comprehensive and robust,’ naming the lockdown restrictions as ‘aggressive but vital’ for containing the spread and developing necessary healthcare infrastructure. The Oxford COVID-19 Government Response Tracker (OxCGRT) appreciated the government’s rapid and stringent actions emphasing on emergency policy-making, emergency investment in health care, fiscal stimulus, investment in vaccine and drug R&D.As matter of fact, India was given a score of 100 for the sudden strict response. Michael Ryan, the chief executive director of the WHO’s health emergencies programme stated that India booms with tremendous capacity to deal with outbreaks owing to its previous experience in eradicating smallpox and polio[7–13]. India on the other hand continues to battle its walk as a country with lowest fatality rates around the globe. This achievement is a combined result of effective application of containment strategy along with house-to-house surveys, quick and aggressive testing and consistent clinical management protocols based on complete Standard of Care approach which ensured that hospitals were left relieved with supervised home isolation for the asymptomatic patients

India’s case fatality rate is one among the lowest in the world at 2.41% as of 23 July and is steadily declining. Six cities were worsely affected in the country – Mumbai, Delhi, Ahmedabad, Chennai, Pune and Kolkata[14]. Lakshadweep is the only region which has not reported a case. India’s recovery rate positions at 63.18% as on 23 July 2020. On 10th June, India’s recoveries surpassed active cases for the first time [19–21]. In this paper an exploratory data analysis and regression analysis is conducted for the cases in India by considering number of cases, testing done, Case Fatality ratio, Number of deaths, change in visits stringency index and measures taken from January to July 2020.

## 2. Data and Methods

Data for this analysis were collected from various sources. The Government of India bulletins publishes the daily details of the condition of Covid-19 situation. Apart from that author uses data from World Health Organization (WHO), ‘Our World in Data’, John Hopkins Corona Resource centre[15–17]. Tweets and Facebook corona updates of Health minister were also used for the study. Media reports around the world were also a source of data. All these sources were used to collect data. MATLAB 2020a is used for the exploratory data analysis. The raw data was cleaned using the software. The platform used for the analysis is Windows 7(64 bit). The regression analysis is done using R. All the data collected and the code generated in MATLAB and R is made available in Github (https://github.com/callmejhe1/Covid-19-India). Following equations were used for the analysis. Linear regression is conducted using the equations

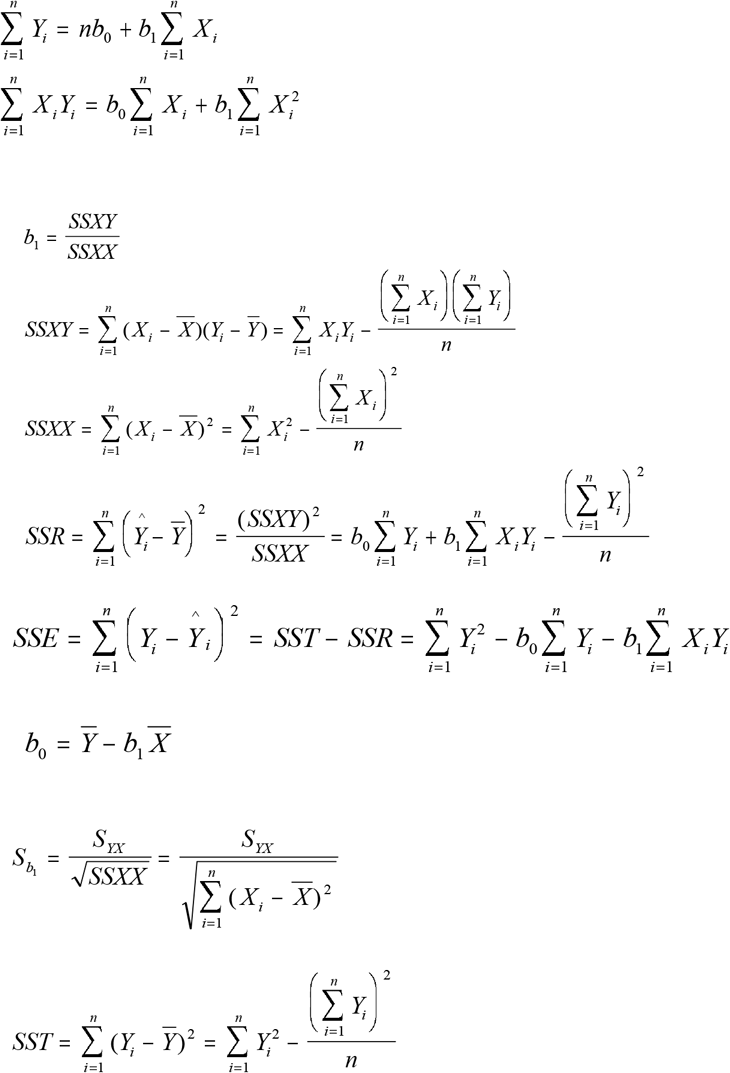

### 2.1 Measures Taken by the country

The outbreak was pronounced as epidemic in many states and union territories, where Epidemic Diseases Act, 1897 have been implemented, leading to the closure of educational and commercial institutions. All tourist transit visas were temporarily suspended, as most of the confirmed cases were mainly from outside[18]. State governments took various methods to contain the spread of the virus inside their local bodies.

In preparation to tackle the pandemic cases various measures and decisions were taken by the state governments as well as Central governments. Throughout the world, the impact of COVID-19 pandemic has been extraordinary, and industries, Small and medium enterprises (SME) in specific, are stressed to maintain and keep up their trade. The government announced intensive measures to ease the affected businesses and taxpayers. The government has asked employers not to dismiss their employees or decrease salaries during the pandemic phase. Additionally, the finance ministry has been immensely working on an economic package to mitigate the impact of coronavirus on the Indian economy. the government announced a complete waiver of interest/late fee for furnishing of returns (GSTR 3B) for taxpayers. The government decided about the basic custom duty (BCD) and Health Cess shall not be charged on import of medical goods. The import of raw materials required for production of these goods has also been granted exemption. The government had introduced a special refund and drawback disposal drive for priority processing and disposal of all pending refund and drawback claims. The measures taken by the government during the pandemic worth appreciation. It provided the much-needed assistance to the industries and eases trade at large.

## 3. Results and Discussion

### 3.1 New Covid-19 Cases

The first case of Covid-19 in India was reported on 30 January in Kerala. There were only three cases by February first week. All were student returnees from China. Twenty two new cases were reported on March 4, 2020. The new cases reported from January to July 2020 are shown in the figure 1. With the cases increasing in the country, nationwide lockdown was implemented on March 25, 2020 for 21 days period which was extended tille May 31,2020. From June first week the unlock process started in three phases. The number of cases were found to be increasing. During may first week, the interstate and inter country travel started which agin increased the number of cases. The number of cases crossed 100000 on May 19.200000 and 1000000 confirmned cases marks were crossed on June 3 and July 2020 respectively. The figure 2 shows the deaths and new cases reported in India during the period.

**Figure 1.**
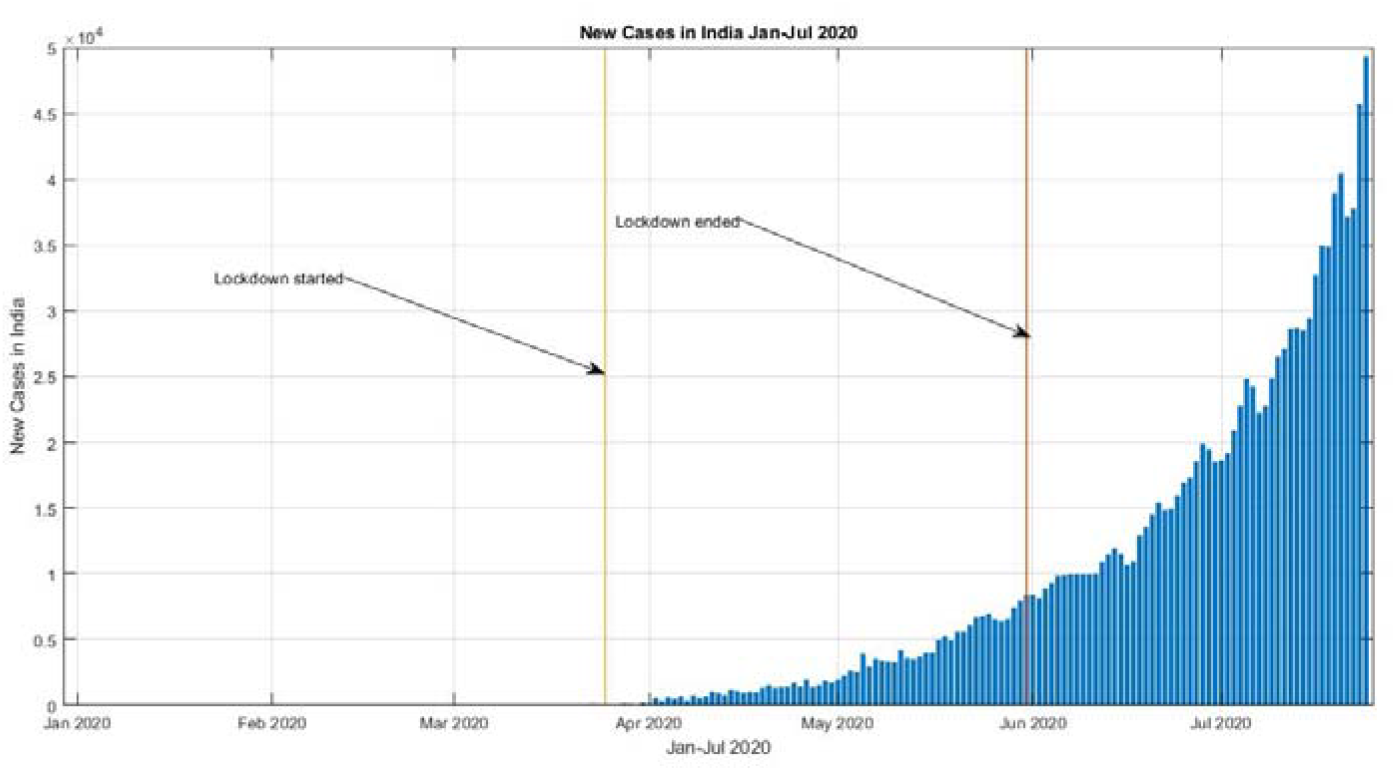
New cases in India reported from Jan – Jul 2020

**Figure 2.**
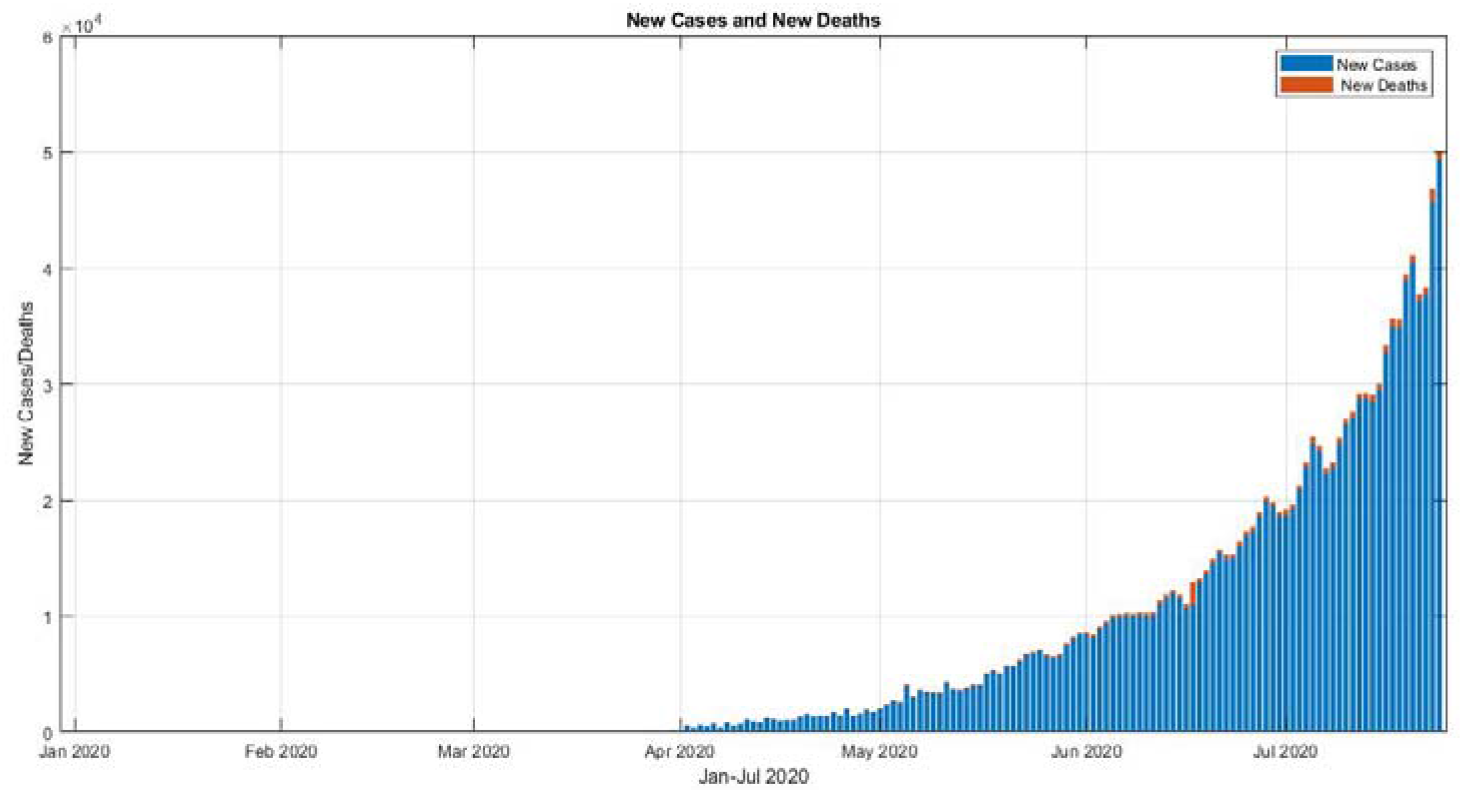
New cases and deaths reported during Jan – Jul 2020

Case fatality ratio(CFR) of India is found to be falling progressively and is 2.25% in July end. This CFR is lowest in the world. Figure 2 represents the stacked chart representing the new cases reported and new deaths due to Covid-19. The CFR was at 3.33% during mid-June 2020. On July 24, the number of cases reached near 15 lakhs. The worst affected state in India is Maharashtra and Tamilnadu. Tamilnadu saw a slow spread of the virus. Andra Pradesh, Delhi and Karnataka had crossed 1 lakh cases.

### 3.2 Testing Strategy

Numbers of testing were initially in a slow phase but gained momentum afterwards. The figure 3 shows the number of testing, cases confirmed and total deaths during the period. Cluster-containment strategy along with breaking the chain of transmission policy was implemented. Scientists at the National Institute of Virology (NIV) isolated a strain of the novel Covid-19 virus. Thus, India became the fifth country to effectively attain a pure sample of the virus after China, Japan, Thailand and the US. Indian council of medical Research mentioned the extraction of pure sample as a step towards developing drugs and vaccines. NIV introduced ELISA antibody test kit which is capable of running 90 samples in a time span of 2.5 hours. During the initial stage person having travel history to 12 designated countries and person coming contact with affected person were tested. All the pneumonia cases regardless of travel history and contact were started to test from March 20. People showing symptoms in the hotspot and containment areas were started to test as per the guidelines by the ICMR. Apart from these, on March 20, accredited private labs were also allowed to conduct the tests as per the guidelines by the Health ministry. With the 65 laboratories of Department of health research and ICMR (DHR-ICMR), test for community spread began on March 15, 2020 from random samples of patients without any travel history.

**Figure 3.**
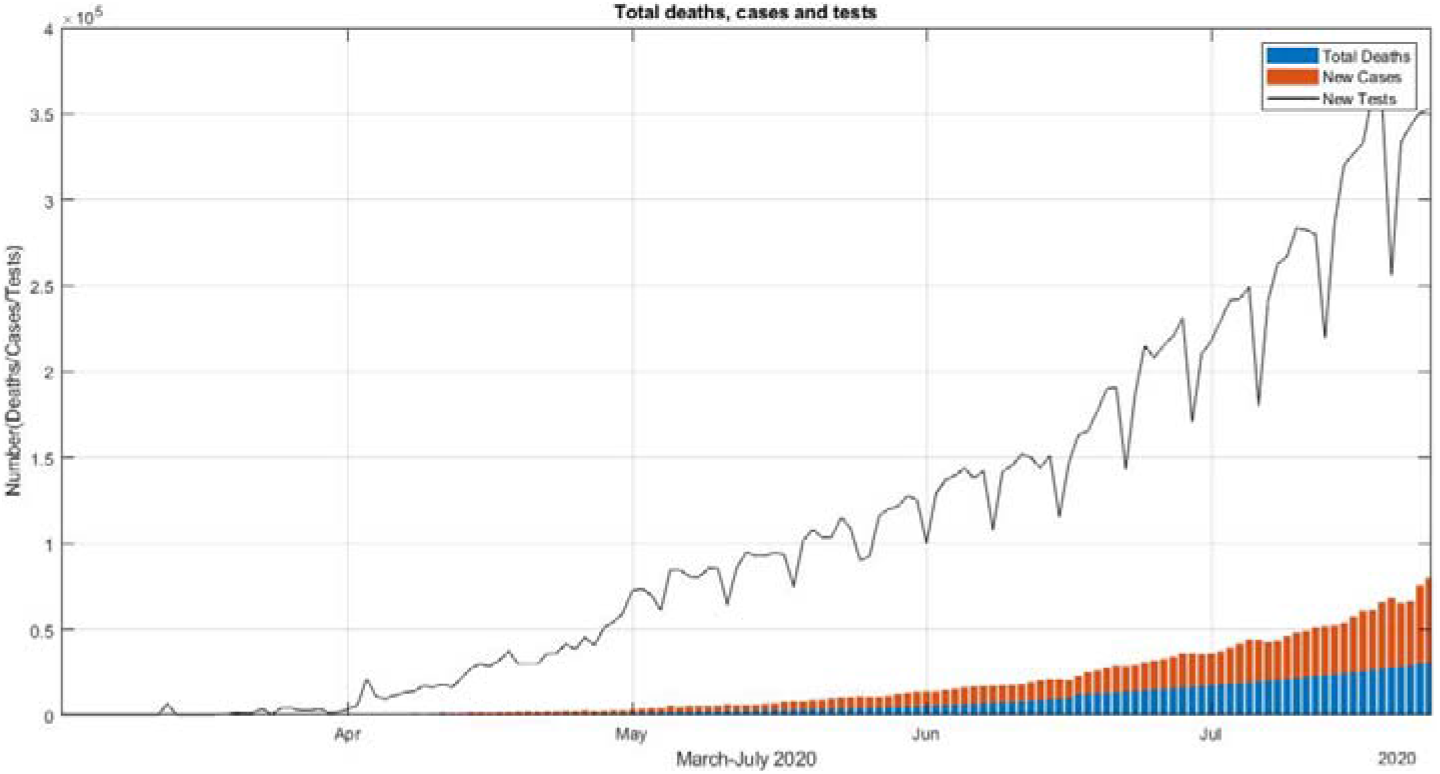
Test conducted deaths and cases repoted

Cumulative tests per million and the cases reported per million is shown the figure 4. In order to develop Covid vaccine completely in India ICMR partnered with Bharath Biotech. 30 candidates were on the road to find vaccine in India. And many were in the pre-clinical stage. During August-October, four vaccines are expected to enter fully clinical trials. During the middle of July 2020, Zydus Cadila had started with human trials for its developed vaccine.

**Figure 4.**
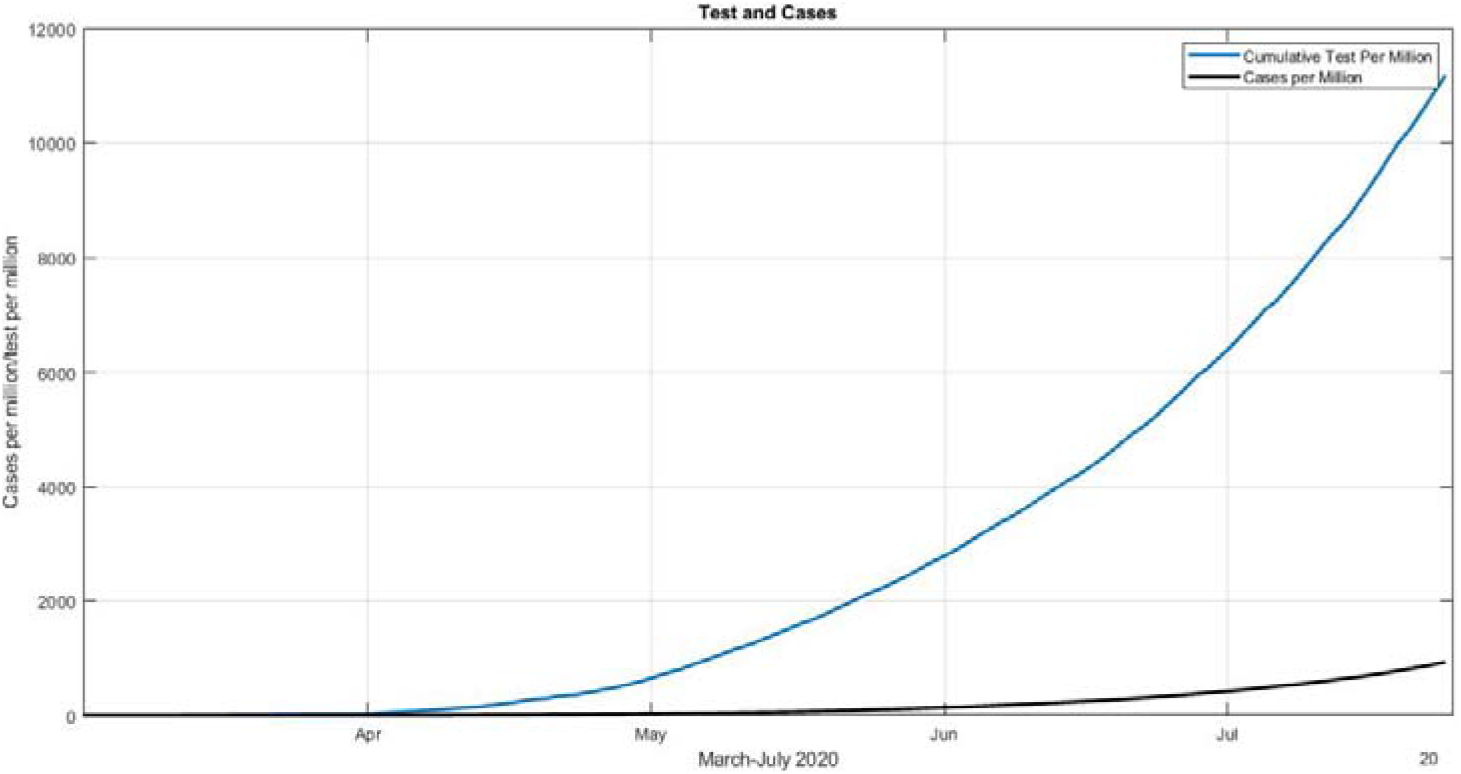
Cumulative test per million and cases per million

### 3.3 Case Fatality Rate and Test per Million

Case fatality ratio (CFR) is the ratio of Confirmed deaths and confirmed cases. The CFR and Test per million is shown in the figure 5. Case fatality ratio(CFR) of India is found to be falling progressively and is 2.25% in July end. This CFR is lowest in the world. CFR of mid-June was 3.33%. Continuous increase in the recovery cases were seen due to the effective measures taken by the centre and state. The recovery rate increased sharply to 64% in July end with respect to 53% in the mid-June 2020.

**Figure 5.**
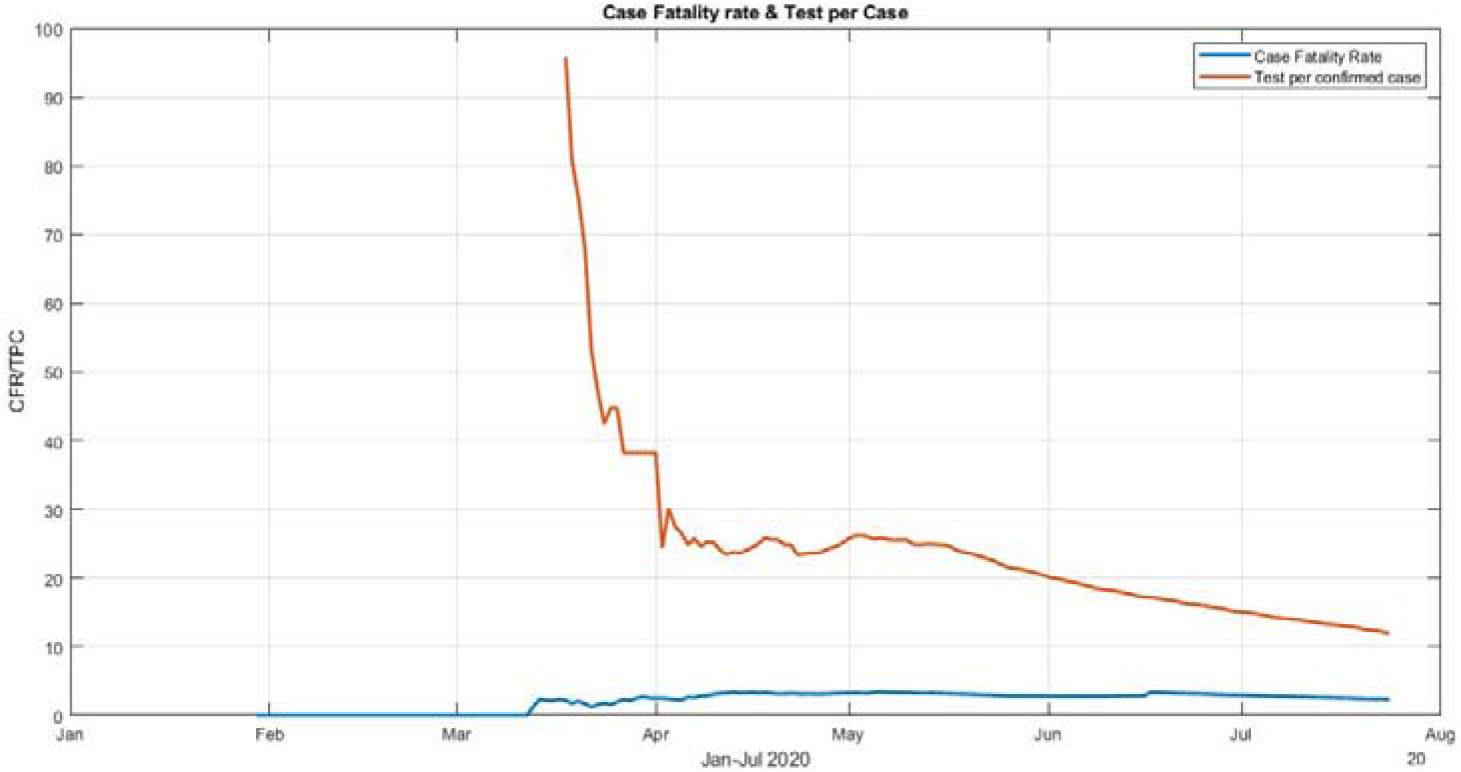
Case Fatality ration and Test per Case

### 3.4 Stringency Index

Stringency index is a score assigned to countries by University of Oxford researchers about the measures taken to fight against the pandemic. The higher score means stricter the country’s measures were. The figure 6 shows the stringency Index during the period. It can be seen that the stringency Index becomes 100 during the lockdown period. The World Health Organization (WHO) and United Nations (UN) have applauded response and action taken by India as ‘comprehensive and robust’. Termed lockdown restriction as ‘aggressive but vital’. The Oxford Covid-19 Government response Tracker (OxCGRT) marked India’s actions to fight against the virus.

**Figure 6.**
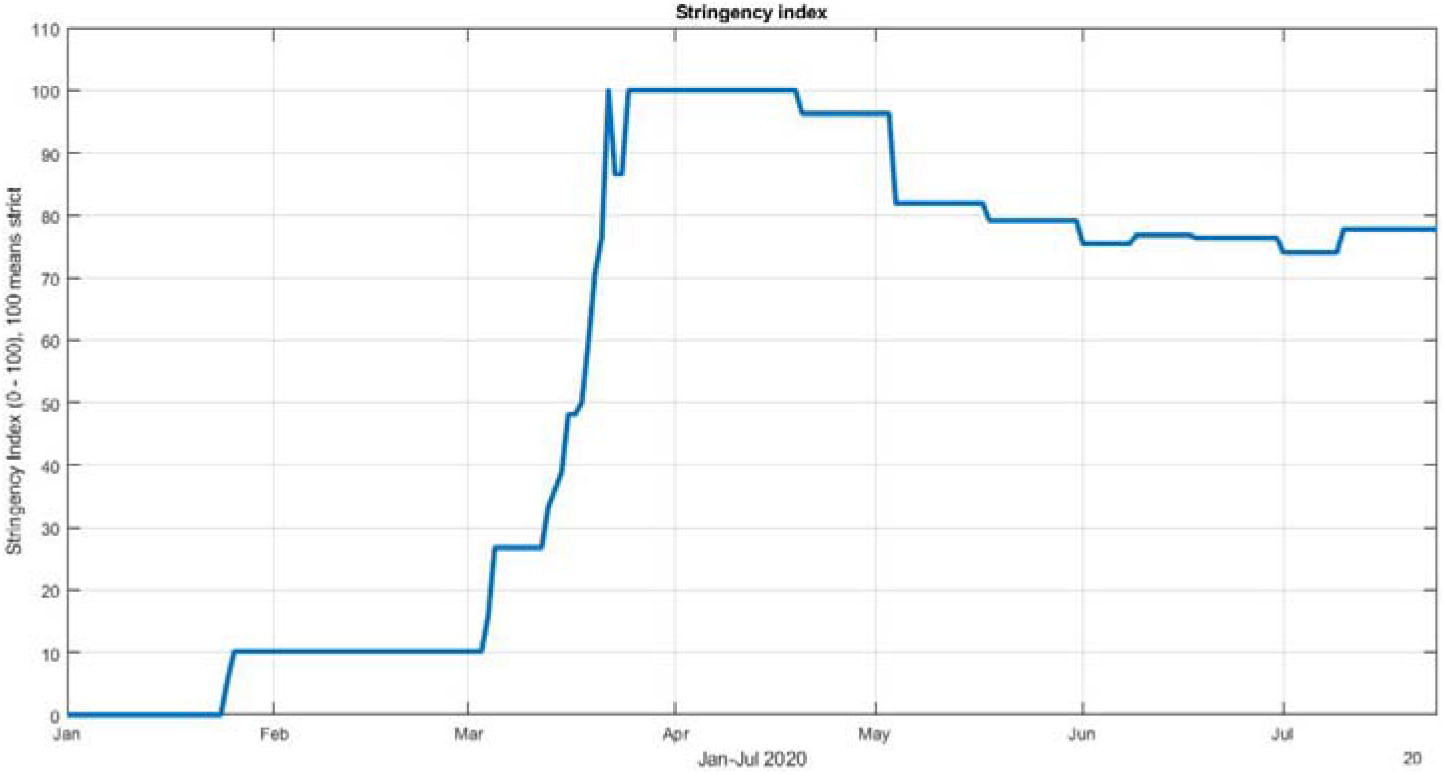
Stringency Index

### 3.5 New test and new cases model

The new cases and new tests done were modelled in a prediction model caret package in R. The code and the data is shown in the Github repository. The correlation between the new cases and new tests is found to be 0.9608, meaning strong relation. Training and testing set were developed. The scatter plot matrix for the new cases, new tests and new deaths is shown in the figure 8a. Linear relationship can be visible from the figure. Linear regression model can also be seen in the figure 8b, 8c and 8d with RMSE = 25120.85, Rsquared = 0.9443099, MAE = 20319.91. The predicted model after the linear regression is shown in the figure 9.

**Figure 7.**
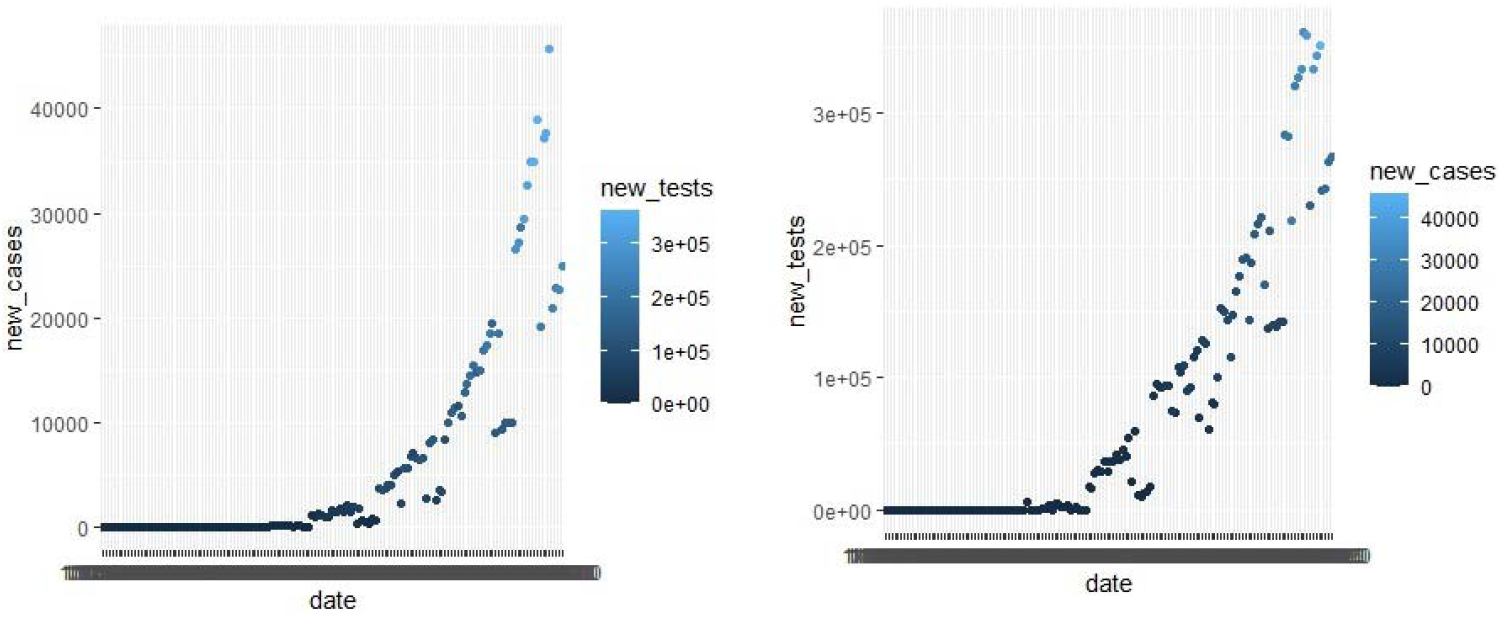
New cases and new tests for modeling

**Figure 8.**
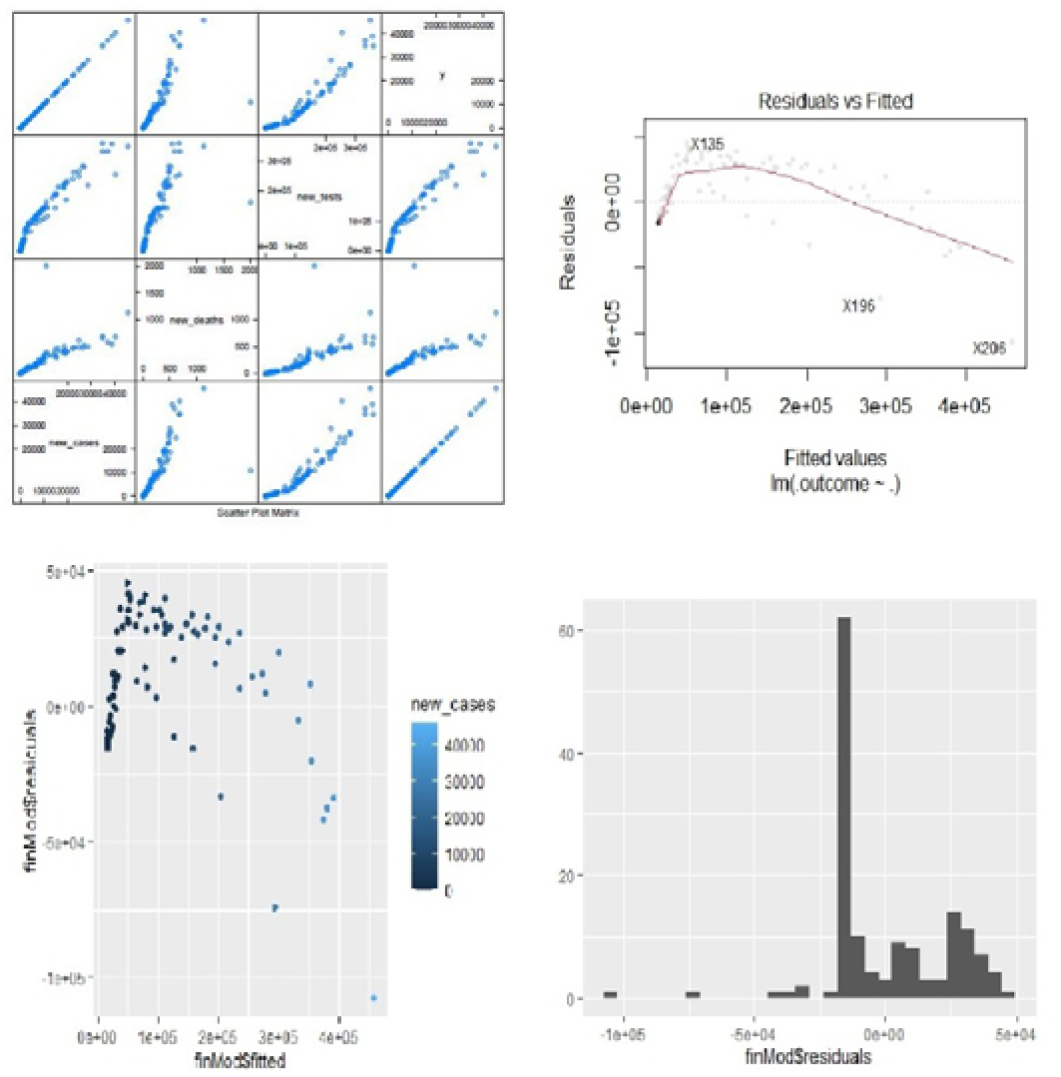
Scatter plot matrix and linear regression fit models

**Figure 9.**
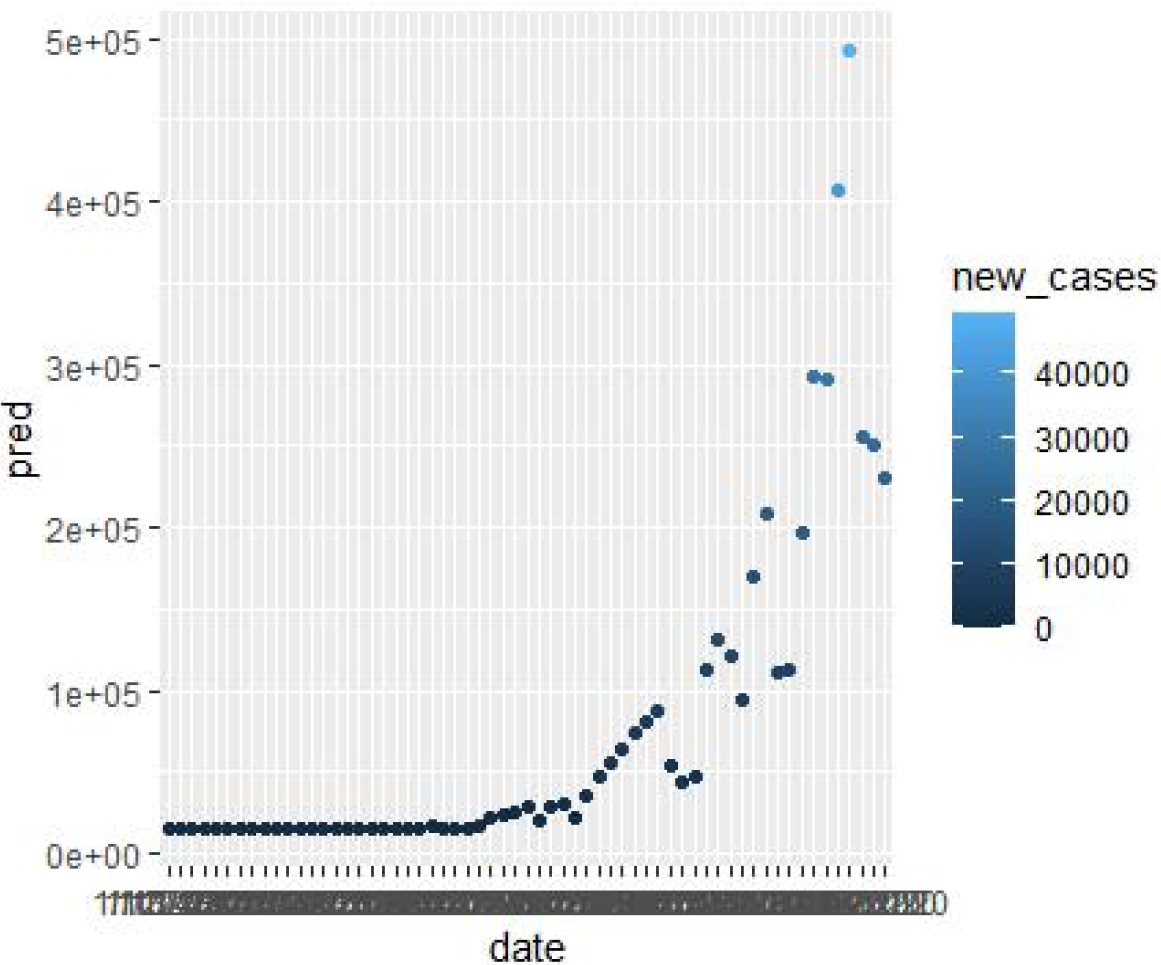
The prediction fit model

### 3.6 Change in visitors

Due to the Covid-19 pandemic many restrictions were imposed in the country. As a result the movement of citizens were also affected. The figure 10 shows the change in visitors at retail and recreation, grocery, pharmaceutical stores, parks, residential, transit station and workplaces. During the period of the pandemic. The zero percentage line represents the normal visits. The parallel plot for the Covid-19 pandemic is shown in the figure 11. Variation of all the elements can be noted from the figure.

**Figure 10.**
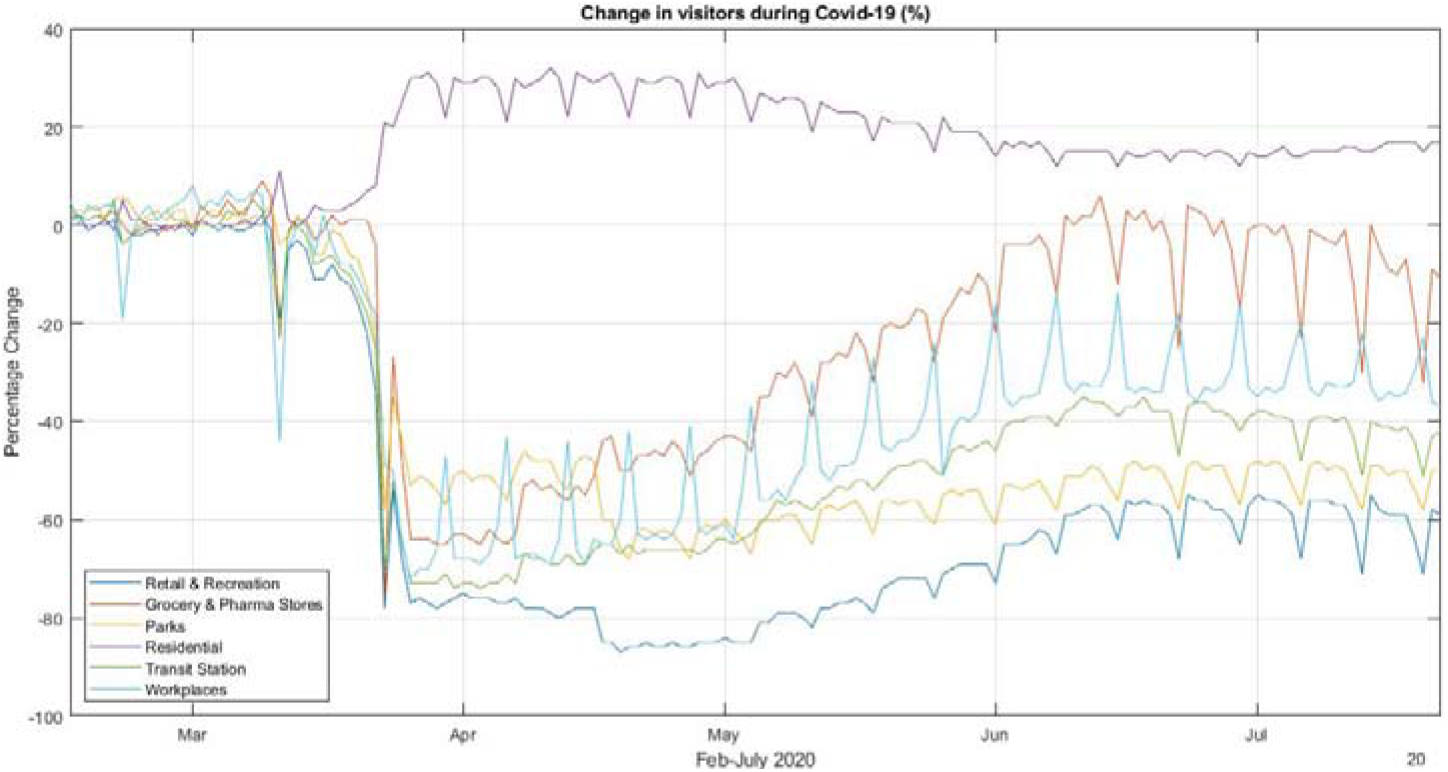
Change in visitors during the pandemic

**Figure 11.**
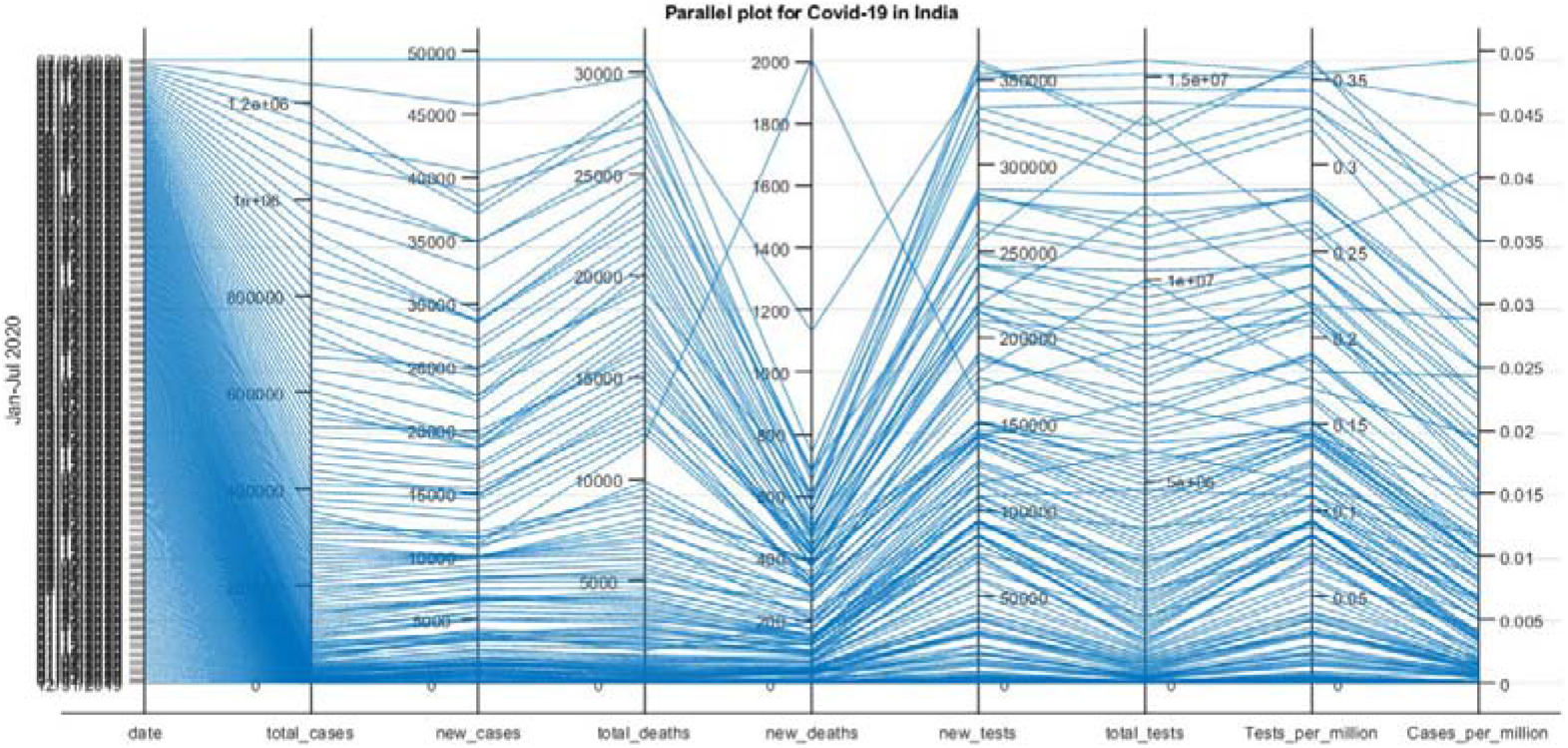
parallel plot showing various element during the period

## 4. Conclusion

An exploratory data analysis was conducted on the Covid-19 cases in India. Data were collected from various sources for the period of January to July 2020. Data was cleaned and arranged in order to do the analysis. MATLAB 2019a and R were used for the analysis. India took stringent measures for fighting against the pandemic. Countrywide lockdown, campaigns, contact tracing, containment zones were introduced for the same. The Oxford Covid-19 Government response Tracker (OxCGRT) marked India’s actions to fight against the virus and gave a score of 100. Case fatality ratio (CFR) of India is found to be falling progressively and is 2.25% in July end. This CFR is lowest in the world. The CFR was at 3.33% during mid-June 2020. The confirmed cases in India crossed 15 lakhs by July end. Number of tests and laboratories were increased progressively. The recovery rate is also found to be climbing up. The visual exploratory data analysis done in this manuscript gives a detail view of the Covid-19 cases in India.

## Data Availability

The data used for the study is made available in the link given below https://github.com/callmejhe1/Covid-19-India

## Acknowledgments

The authors would like to thank the World Health Organization, ICMR, Our World in Data, Johns Hopkins University for open sourcing their dataset. Authors would also like to thank the Health Department, India for their timely daily bulletin reports on Covid-19 cases.

## Ethical Approval

This study does not contain any studies with human participants or animals performed by any of the authors.

## Informed consent

This study does not contain any studies with human participants.

